# DEFINING CANDIDATE PARKINSON’S DISEASE GENES THROUGH THE ANALYSIS OF GENOME-WIDE HOMOZYGOSITY

**DOI:** 10.1101/2020.11.23.20235671

**Authors:** Steven J Lubbe, Yvette C. Wong, Bernabe Bustos, Soojin Kim, Jana Vandrovcova, Brendan P Norman, Manuela Tan, David Murphy, Camille B Carroll, Donald Grosset, Mina Ryten, Juan A. Botía, Valentina Escott-Price, Nigel M Williams, Dimitri Krainc, Huw R Morris, for International Parkinson’s Disease Genomics Consortium

## Abstract

Early-onset Parkinson’s disease (EOPD) can be caused by biallelic mutations in *PRKN, DJ1* and *PINK1*. However, while the identification of novel genes is becoming increasingly challenging, new insights into EOPD genetics have important relevance for understanding the pathways driving disease pathogenesis. Here, using extended runs of homozygosity (ROH) >8Mb as a marker for possible autosomal recessive inheritance, we identified 90 EOPD patients with extended ROH. Investigating rare, damaging homozygous variants to identify candidate genes for EOPD, 81 genes were prioritised. Through the assessment of biallelic (homozygous and compound heterozygous) variant frequencies in cases and controls from three independent cohorts totalling 3,381 PD patients and 2,463 controls, we identified two biallelic *MIEF1* variant carriers among EOPD patients. We further investigated the role of disease-associated variants in *MIEF1* which encodes for MID51, an outer mitochondrial membrane protein, and found that putative EOPD-associated variants in MID51 preferentially disrupted its oligomerization state. These findings provide further support for the role of mitochondrial dysfunction in the development of PD. Together, we have used genome-wide homozygosity mapping to identify potential EOPD genes, and future studies incorporating expanded datasets and further functional analyses will help to determine their roles in disease aetiology.

## INTRODUCTION

Parkinson’s disease is the second most common neurodegenerative disorder, with around 1% of Parkinson’s cases having an age at onset below 40 years characterized as early-onset Parkinson’s disease (EOPD) (Wickremaratchi et al., 2009). There are several Mendelian forms of EOPD (Lubbe and Morris, 2014) with mutations in autosomal recessive (AR) genes (*PRKN, DJ1* and *PINK1*) common in EOPD. Pathogenic biallelic mutations in these genes are more common in cases with the youngest ages of onset, as well as those with a positive family history or parental consanguinity (Kilarski et al., 2012). While progress has been made in identifying additional genes linked to EOPD, currently only between 5-12% of EOPD cases have an identified single-gene cause, including substantial numbers of familial PD cases (Kilarski et al., 2012; Zhao et al., 2020). PD kindred analysis shows a greatly increased sibling as compared to parental recurrence risk in EOPD (Marder et al., 2003). Together, this suggests that there are likely to be additional AR PD genes that have not yet been identified.

Homozygosity (HMZ) mapping in consanguineous families in conjunction with whole-exome sequencing (WES) data is a powerful approach that has successfully led to the recent identification of the AR *DNAJC6* (Edvardson et al., 2012) and *VPS13C* (Lesage et al., 2016) genes in autosomal recessive parkinsonism. Since their discovery, studies have attempted to identify additional *DNAJC6* (Elsayed et al., 2016; Köroğlu et al., 2013; Olgiati et al., 2016) and *VPS13C* (Darvish et al., 2018; Schormair et al., 2018) mutation carriers, and have highlighted that these genes are very rare causes of EOPD – estimated to account for 0.8% (Olgiati et al., 2016) and 0.2% (Lesage et al., 2016) of EOPD cases, respectively. This suggests that it is likely that new AR genes will be very rare and difficult to identify using population-based approaches.

Using extended runs of homozygosity (ROH) as a marker of genomic HMZ, we found a significant excess of individuals with ROH >8Mb in 1,445 EOPD cases compared to 6,987 controls (Simón-Sánchez et al., 2012). This excess, which was independent of known AR EOPD genes, related to 71 EOPD cases who were found to have an earlier age at onset and mean inbreeding coefficients 10-times higher than the remaining EOPD cases and controls, suggesting increased relatedness. We hypothesised that EOPD cases with extended ROH harbour new AR PD genes and report WES analysis in 90 homozygous EOPD cases identified from two large WES independent cohorts totalling 1,736 EOPD and 1,413 controls. Following the assessment of the top candidates in an additional replication cohort (Accelerating Medicines Partnership: Parkinson’s Disease (AMP-PD, https://amp-pd.org) cohort totalling 1,647 PD cases and 1,050 controls), we provide functional evidence for a role of biallelic *MIEF1* variants in EOPD pathogenesis.

## RESULTS

### Identification of novel candidate autosomal recessive PD genes using large-scale genetic cohorts

Using cohorts pre-screened for known PD- or Parkinsonism-associated mutations (*e.g. LRRK2, PRKN, SNCA, PINK1, DJ1, ATP13A2, FBXO7, VPS35, VPS13C, DNAJC6, SYNJ1, GBA, GCH1 etc*.), we identified 90 EOPD patients with extended ROHs >8Mb, in which we sought novel disease associated AR mutations. From WES data on these 90 patients, we identified ≥1 rare (MAF<0.01) homozygous loss of function variants (nonsense, splice-site or frameshift insertion/deletion) or nonsynonymous variants with CADD phred score ≥15 (together termed deleterious nsSNVs) (Supplementary Table 1) in 81 genes.

In these 81 genes we first compared the rate of biallelic deleterious variant carriers in EOPD cases as compared to controls in two independent cohorts totalling 1,734 EOPD cases and 1,413 controls. We identified 19 genes in which there were more frequent biallelic variant carriers in cases as compared to controls, but no gene reached statistical significance (Supplementary Table 1). Seven genes (*GPRIN2, GXYLT1, HSH2D, HLA-DQA1, SRA1, WWTR1*, and *ZNF492*) were excluded due to a high observed carrier frequency (>1%) of prioritised variants within the replication cohorts. Although not significantly different, three genes were found to have at least twice as many carriers in cases compared to controls: *DYSF*, OR=2.06, 95% CI: 0.24-17.72; *MARF1*, OR=3.74, 95% CI: 0.34-41.40; *SIRPB1*, OR=4.57, 95% CI: 0.59-35.46. However, the *SIRPB1* p.V131Gfs*8 and c.432C>T splice variants are 39bp apart (Supplementary Table 2), they usually appear together, and are likely on a single haplotype – therefore *SIRPB1* was excluded from further analyses. We then assessed the remaining 11 genes in the AMP-PD cohort (PD=1,647, controls=1,050) to identify additional carriers of rare biallelic deleterious nsSNVs in cases (Supplementary Table 3). *CYP4B1* and *DYSF* were found to have more carriers in cases (n=3 for each gene) compared to controls (n=1; OR=1.91, 95% CI: 0.15-100.57, P=0.49), while single cases (and no controls) were observed for *MARF1* and *PLG* (P=0.61 each).

As family members were not available to test for variant segregation, we next assessed the combined counts using sequencing data across the discovery and all three independent replication cohorts totalling 3,471 cases and 2,463 controls (Table 1). Two genes had higher frequencies of biallelic damaging variant carriers in cases versus controls, providing some evidence of potential enrichment in EOPD: *MARF1* (OR=1.91, 95% CI: 0.38-21.60, P=0.29) and *RBM14* (OR=2.13, 95% CI: 0.17-111.84, P=0.45). Three cases with biallelic *PLG* LOF variants were seen (3/3,471, 0.09%), while *MIEF1, SCARF2, SEC24A* and *SYTL3* all harboured biallelic LOF variants in EOPD cases only (2/3,471, 0.06%).

**Table 1:** Assessment of carrier frequency of biallelic rare, damaging variants in Parkinson’s disease cases compared to controls across three independent combined cohorts totalling 3,471 cases and 2,463 controls.

| Gene | Cases (n=3,471) |  | Controls (n=2,463) |  | Fisher's exact |  |
| --- | --- | --- | --- | --- | --- | --- |
|  | Carriers | Freq | Carriers | Freq | P-value | OR (95% CI) |
| *CYP4B1* | 9 | 0.0026 | 5 | 0.0020 | 0.439 | 1.28 (0.38-4.86) |
| *DYSF* | 9 | 0.0026 | 7 | 0.0028 | 0.523 | 0.91 (0.30-2.89) |
| *EFCAB6* | 12 | 0.0035 | 8 | 0.0032 | 0.541 | 1.06 (0.40-4.01) |
| *FMO2* | 4 | 0.0012 | 4 | 0.0016 | 0.441 | 0.71 (0.13-3.81) |
| *MARF1* | 6 | 0.0017 | 2 | 0.0008 | 0.285 | 2.13 (0.38-21.60) |
| *MIEF1* | 2 | 0.0006 | 0 | 0 | 0.342 | - |
| *PLG* | 3 | 0.0009 | 0 | 0 | 0.200 | - |
| *RBM14* | 3 | 0.0009 | 1 | 0.0004 | 0.449 | 2.13 (0.17-111.84) |
| *SCARF2* | 2 | 0.0006 | 0 | 0 | 0.342 | - |
| *SEC24A* | 2 | 0.0006 | 0 | 0 | 0.342 | - |
| *SYTL3* | 2 | 0.0006 | 0 | 0 | 0.342 | - |

Damaging variants are defined as all loss of function (stop gains, frameshift insertions/deletions, splicing) variants and nonsynonymous variants predicted damaging by CADD (phred score >15). The combined data reflects the total samples across the discovery cohort (PD=90), Accelerating Medicines Partnership: Parkinson's Disease (AMP-PD) cohort (PD=1,647, controls=1,050), International Parkinson's Disease Genomics Consortium (IPDGC) cohort (PD=1,170, controls=482), and the University College London (UCL) cohort (PD=564, controls=931). Uncorrected P-values are shown. *GPRIN2*, *GXYLT1*, *HLA-DQA1*, *HSH2D*, *SRA1*, *WWTR1*, and *ZNF492* are not shown due to their exclusion as candidates.
Key: CADD, Combined Annotation Dependent Depletion score (https://cadd.gs.washington.edu/); biallelic (homozygous and compound heterozygous) variant carriers; CI, confidence intervals; Freq, frequency; n, number; OR, odds ratio; %, percent.

Observing candidate variants identified in the discovery cohort, within independent replication cases, allows for genes and variants to be further prioritised (Supplementary Table 4). The *MIEF1* discovery case harboured a homozygous p.R169W (rs2232088) potentially deleterious nsSNV, and the single replication case carried heterozygous p.A53V (rs373246269) and p.R169W deleterious nsSNVs. Combined with the observed increased frequency of variants, we prioritised *MIEF1, MARF1, RBM14*, and *SEC24A* as potential EOPD candidate genes. The *MARF1* and *RBM14* discovery variants were not confirmed by Sanger sequencing in available DNA samples and are potential sequencing artefacts. The *SEC24A* discovery variant was confirmed by Sanger sequencing; however not all of the replication variants were confirmed. Thus, we focused on the candidate *MIEF1* variants which we directly confirmed by Sanger sequencing, and subsequently explored their potential damaging role in misregulating *MIEF1*.

### PD-linked MIEF1 variants preferentially disrupt its oligomerisation

*MIEF1* encodes Mitochondrial dynamics protein of 51 kDa (MID51), an outer mitochondrial membrane protein which regulates mitochondrial fission dynamics (Palmer et al., 2011; Zhao et al., 2011) and which oligomerises to regulate its function (Losón et al., 2014; Zhao et al., 2011). Homozygous or compound heterozygous mutations in *MIEF1* have not been previously linked to any human disease. We first examined whether the identified *MIEF1* variants (p.R169W and p.A53V) prevented its mitochondrial localisation using high spatial and temporal confocal microscopy in live cells. Expression of MID51 (wild-type, WT) robustly localised to mitochondria (Figure 1a,b), consistent with its reported localisation on the outer mitochondrial membrane (Palmer et al., 2011; Zhao et al., 2011). We found that the p.R169W and p.A53V variants did not disrupt this localisation as MID51 maintained its localisation to mitochondria in live cells for both variants (Figure 1a,b).

**Figure 1:**
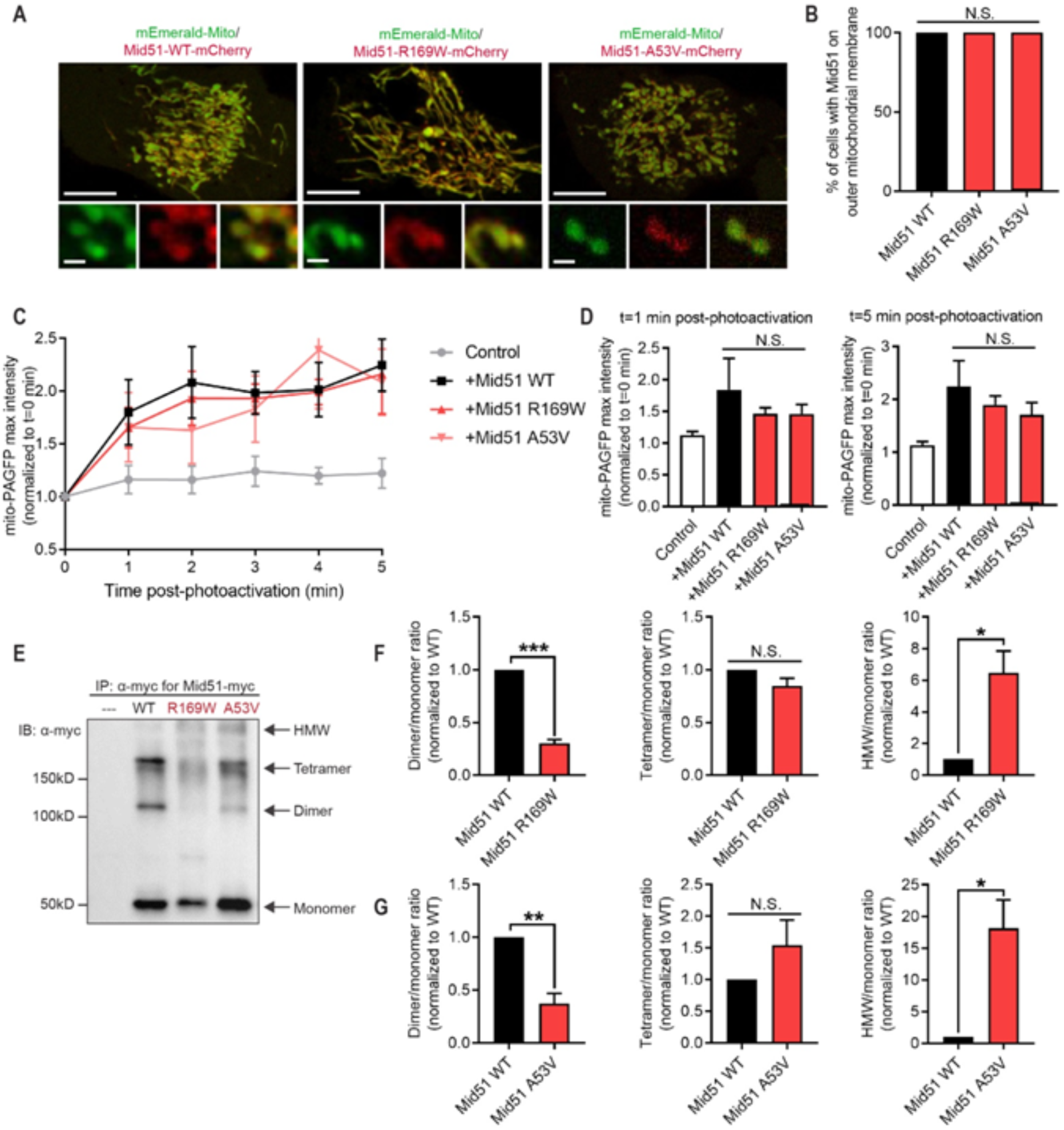
Parkinson’s disease-linked *MIEF1* variants preferentially disrupt MID51 oligomerisation. (A-B) Representative live-cell confocal images and quantification of mCherry-tagged MID51 (WT; p.R169W; p.A53V) showing MID51 localisation to mitochondria (mEmerald-Mito) in HeLa cells. (n=100 cells from N=3 experiments per condition). Scale bar, 10µm (inset, 1µm). (C-D) Representative traces and analysis of mito-PAGFP fluorescence intensity in distal region (10µm from site of mitochondrial photoactivation) showing similar mitochondrial fusion rates between MID51 (WT) and MID51 variant p.R169W and p.A53V. (n=9 cells (control); n=10 cells (WT); n=13 cells (p.R169W); n=13 cells (p.A53V) from n=3 experiments per condition). (E-G) Representative immunoblot (IB: α-myc) and quantification of Mid51 oligomerisation from immunoprecipitation (IP: α-myc) of myc-tagged MID51 (WT; p.R169W; p.A53V) and control (---), showing defective oligomerisation (decreased dimer and increased high molecular weight (HMW) species) in both MID51 variants (p.R169W; p.A53V) compared to wild-type MID51 (WT). Data are means ± s.e.m. (\**P* < 0.05; \*\**P* < 0.01; \*\*\**P* < 0.001; N.S.=not significant; ANOVA with Tukey’s post-hoc test (B,D) or unpaired two-tailed *t*-test (F,G)).

As MID51 is a known regulator of mitochondrial dynamics, mitochondrial fission/fusion dynamics were further assessed to examine whether they were disrupted by the identified variants. WT MID51 expression has previously been shown to inhibit mitochondrial fission, resulting in increased mitochondrial fusion events (Zhao et al., 2011). To examine this, WT and MID51 variants were expressed and the rate of mitochondrial fusion events in live cells was analysed using a photo-activatable mitochondrial matrix probe (mito-PAGFP). A subpopulation of mitochondria were selectively photoactivated, and the rate of fusion events with neighbouring mitochondria were analysed, measured as an increase over time in fluorescence in neighbouring mitochondria. Consistent with previous reports, WT MID51 expression increased the rate of fusion events (Figure 1c,d) compared to control cells. However, the p.R169W and p.A53V variants were also able to increase the rate of fusion events (Figure 1c,d), and they were not significantly different from WT MID51, suggesting that expression of the MID51 variants does not inhibit the protein’s capacity to promote mitochondrial fusion events.

Finally, as MID51 has been shown to oligomerise to regulate its function (Losón et al., 2014; Zhao et al., 2011), we assessed whether the *MIEF1* variants disrupted oligomerisation. To test this, MID51 oligomerisation state was analysed following immunoprecipitation (IP) for myc-tagged MID51 WT, p.R169W and p.A53V. While MID51 WT formed distinct monomer, dimer and tetramer bands, there were few high molecular weight (HMW) species (Figure 1e). In contrast, both p.R169W (Figure 1f) and p.A53V (Figure 1g) showed significantly decreased dimer formation (Figure 1e-g) but increased HMW formation (Figure 1e-g), resulting in decreased dimer/HMW ratios for both p.R169W and p.A53V compared to Mid51 (WT) (*\*\*\*p<0.001*, MID51 (p.R169W) = 0.048 ± 0.006; *\*\*\*p<0.001*, MID51 (p.A53V) = 0.028 ± 0.002; values normalised to MID51 (WT)).

Together, these data show that the identified *MIEF1* variants do not prevent MID51 localisation to the outer mitochondrial membrane or its ability to promote mitochondrial fusion, but preferentially shift its dimer formation towards HMW oligomers resulting in a disruption of its oligomerisation state.

## DISCUSSION

Extended ROHs (>8Mb) were used as markers for individuals most likely to harbour homozygous AR variants accounting for their disease status. From a combined discovery cohort of 90 HMZ (ROH>8Mb) samples, genes harbouring homozygous rare damaging variants were prioritised for replication in three independent case-control replication series totalling 3,471 EOPD cases and 2,463 controls. Eleven genes had suggestive evidence of enrichment in EOPD cases compared to controls. A combined analysis of the discovery and all three cohorts (Table 1) identified evidence of excess carriers in seven genes, as suggested by higher carrier frequencies in cases compared to controls, albeit non-statistically significant. Rare deleterious nsSNVs in the discovery cohort and one of the replication cohorts were seen for *MIEF1* (p.R169W), *MARF1* (p.V865L), *RBM14* (p.A493L), and *SEC24A* (p.P1018L). These genes represent very rare potential candidates for further investigation in the aetiology of EOPD. Following variant validation, the candidate gene *MIEF1* was taken forward for functional assessment to explore whether these deleterious nsSNV might contribute to abnormal cell biology.

*MIEF1* encodes for the protein MID51, which localises to the outer mitochondrial membrane and is an important regulator of mitochondrial dynamics (Kalia et al., 2018; Koirala et al., 2013; Losón et al., 2013; Palmer et al., 2013, 2011; Zhao et al., 2011). Changes in mitochondrial dynamics and mitophagy appear to be key mechanisms in EOPD. Well established pathogenic mutations in *PRKN* and *PINK1* disrupt mitophagy and lead to abnormal mitochondrial dynamics (Dagda et al., 2009; Mortiboys et al., 2008). In this study, we show that the *MIEF1* variants (p.R169W and p.A53V) disrupted the MID51 oligomerisation state, resulting in reduced dimers and increased HMW forms, which may be pathological. These are consistent with previous findings that the region containing these residues is required for MID51 dimerization (Zhao et al., 2011), and that MID51 Arg169 is an important residue for its dimerization (Losón et al., 2014). As only homozygous (p.R169W/p.R169W) or compound heterozygous (p.A53V/p.R169W) variants were observed in EOPD, proper oligomerisation of WT MID51 may be sufficient to compensate in heterozygous carriers. In contrast, both *MIEF1* variants we identified, which are outside of MID51’s N-terminal transmembrane domain (Richter et al., 2014; Zhao et al., 2011), did not disrupt protein localisation to the outer mitochondrial membrane. Furthermore, both variants did not prevent its ability to promote mitochondrial fusion by inhibiting fission. This is consistent with the fact that neither residue is directly located in regions which regulate DRP1 recruitment (Ma et al., 2019; Richter et al., 2014), the GTPase which drives mitochondrial fission, and the finding that MID51 oligomerisation is not required for DRP1 recruitment (Losón et al., 2014). However, MID51’s oligomerisation state has been proposed to be important for regulating its overall mitochondrial function including promoting the oligomerisation of DRP1 (Losón et al., 2013). As dopaminergic neurons are particularly susceptible to mitochondrial defects (Berthet et al., 2014; Pickrell et al., 2015), it is possible that mitochondrial dysfunction may further contribute to neurodegeneration in *MIEF1*-associated PD patients.

The *MIEF1* deleterious nsNSVs identified here appear to be relatively common in European ancestry individuals (p.R169W, 0.79%). However, homozygous p.R169W carriers are relatively rare with a background frequency of 0.014% (9/64,536) in Europeans. If p.R169W has a role in PD then it is likely that there is incomplete penetrance. Biallelic *PRKN* mutations are the most common cause of AR EOPD (MIM#600116) and are known to have a large range of age of onset. Many different mutations are linked to disease, however not every *PRKN* mutation has equal penetrance (*e.g*. p.R275W) as suggested by varying ages of onset in families and in carriers of different *PRKN* mutations (Pankratz and Foroud, 2007). Incomplete penetrance occurs in autosomal dominant PD, where *LRRK2* p.G2019S has an age-dependent penetrance estimated to be between 25% and 42.5% by age of 80 (Healy et al., 2008; Lee et al., 2017). Reduced penetrance complicates the identification of disease-associated variants and points to possible additional factors at work, such as modifier genes/variants as has recently been shown for *LRRK2* p.G2019S (Iwaki et al., 2020). Replication of these results in additional larger case-control cohorts will be required to accurately assess the role and penetrance of *MIEF1* variants in the pathogenesis of EOPD.

There are a series of limitations in using WES data to try to identify novel Mendelian EOPD genes in large single case-control cohorts, even when these are focussed on young-onset or homozygous disease. Firstly, the remaining genes/mutations may be “private” (Bobbili et al., 2020), affecting a very small number of families. Biallelic mutations in *VPS13C* were recently linked to EOPD and are estimated to account for 0.2% of EOPD cases (Lesage et al., 2016). The IPDGC WES data used here has previously been used to identify and validate *VPS13C* variants linked to EOPD (Jansen et al., 2017; Lesage et al., 2016) where a single compound heterozygous mutation carrier was observed (1/1,148 EOPD; 0.09%). Compared to zero (0/503) controls, this would not return a statistically significant replication result (Fisher’s exact P=0.695) – therefore adequate power to detect or replicate any associations was low.

The discovery of *VPS13C* and other recent AR genes was made possible due to segregation analysis in large multiplex pedigrees (Lesage et al., 2016; Olgiati et al., 2016; Quadri et al., 2013). Although some of the largest PD cohorts available have been used in this study, *MIEF1* remains a candidate EOPD gene until adequate replication in additional larger cohorts is achieved, or further families for segregation can be studied. However, it has been suggested that cohort studies aiming to uncover rare disease-associated variants require in excess of 25,000 cases before adequate power is achieved (Zuk et al., 2014). Secondly, assessing the segregation of candidate variants within families is essential; however, this is difficult in cases where samples have been collected over many years as part of large international consortia. Thirdly, WES has limited coverage and may have a significant false positive rate. We have also not interrogated noncoding variants, copy number variants, structural variants, short tandem repeats *etc*. Using whole-genome sequencing would overcome these issues by allowing for more accurate variant identification and for the assessment of different types of variants. Fourthly, particularly in PD, it may be difficult to disentangle the contribution of polygenic risk and high penetrance gene mutations, and we have previously identified polygenic risk as being important in driving EOPD (Escott-Price et al., 2015; Nalls et al., 2015).

Some of the remaining genes that demonstrated some evidence of enrichment in EOPD can also be considered plausible EOPD candidate genes. SYTL3, which binds to phospholipid membranes in a Ca^2+^-dependent manner (Fukuda, 2002), belongs to a family of peripheral membrane proteins that are characterized by C-terminal tandem C2 domains and are known to play a role in vesicular trafficking (Nalefski and Falke, 1996). The *SCARF2* gene encodes a Ca^2+^-binding scavenger receptor that forms heterodimers with SCARF1 thereby inhibiting SCARF1-mediated endocytosis of chemically modified lipoproteins such as acetylated low-density lipoprotein (Zani et al., 2015). *SCARF2* maps to the 22q11 deletion region which has been associated with increased PD risk (Mok et al., 2016) – the underlying mechanism at this locus has yet to be identified. Plasminogen, encoded for by the *PLG* gene, is the precursor for the enzyme plasmin. Plasmin has been shown to cleave both aggregated and monomeric extracellular SNCA thereby preventing its translocation into neighbouring cells as well as the activation of microglia and astrocytes (Kim et al., 2012). *SEC24A* encodes a coat protein complex II (COPII) subunit known to be involved the selection of proteins for transport to the Golgi, as well as in the derivation of endoplasmic reticulum-derived vesicles to the Golgi apparatus (Bonnon et al., 2010; Mancias and Goldberg, 2008, 2007).

It is becoming increasingly difficult to accurately identify the role of rare mutations in the development of complex diseases like PD. Here we show that the use of samples with extended ROHs and therefore high genomic homozygosity as a discovery set to identify rare homozygous variants in candidate PD genes is a valid sensitive approach. In spite of the apparent lack of statistical power, we provide some evidence to support a role for rare biallelic damaging variants in *MIEF1* in PD. These findings may provide further support for the important roles of mitochondria in the development of PD. However, additional carriers of *MIEF1* LOF variants in EOPD, family-based studies, as well as additional functional/cellular studies are needed before a role for *MIEF1* can be conclusively linked to PD aetiology.

## METHODS AND MATERIALS

### Patients and samples

For candidate gene identification, a discovery/replication approach was used. The discovery cohort comprised 90 EOPD samples from the International Parkinson’s Disease Genomics Consortium exome dataset (IPDGC) (n=28) and University College London (UCL) exomes dataset (n=62). In each case, high HMZ was defined by possession of a ROH ≥8Mb (autosomes only, not overlapping centromeres). The following independent replication cohorts were used: (i) WES data from 1,170 PD cases and 482 controls from the IPDGC, and are described in detail elsewhere (https://pdgenetics.org/resources); (ii) WES data from 564 cases and 931 controls from UCL, which comprise samples collected in the PROBAND/Tracking Parkinson’s project, Parkinson’s families project REC 15/LO/0097 and Molecular Genetic Studies of Neurodegenerative disorders project (Malek et al., 2015); and (iii) the AMP-PD whole-genome sequencing data including 1,647 PD cases and 1,050 healthy controls of European descent from three different cohorts (BioFind; https://biofind.loni.usc.edu/), Parkinson’s Disease Biomarker Program (PDBP; https://pdbp.ninds.nih.gov/), and Parkinson’s Progression Markers Initiative (PPMI; https://www.ppmi-info.org/). Cohort characteristics and quality control procedures are described elsewhere (https://amp-pd.org/whole-genome-data).

### Whole-exome sequencing

Sample libraries were prepared using Illumina, Nimblegen (IPDGC and UCL) or Agilent (UCL) capture kits with paired-end sequencing performed on the Illumina HiSeq2000. All reads were aligned using BWA against the UCSC hg19 reference genome (Li and Durbin, 2009). Variant calling and quality-based filtering were done using GATK (McKenna et al., 2010). All FASTQ files for each cohort were analysed on the same pipeline at the same time to ensure high quality genotypes common to all capture methods used. To ensure data compatibility between capture platforms, variants with >10% missing genotype calls, variants with low coverage (DP<8), variants within segmental duplications and those failing quality filters were excluded.

ANNOVAR (Wang et al., 2010) was used to annotate variants with Combined Annotation Dependent Depletion (CADD) scores to predict the relative impact of observed variants (Rentzsch et al., 2019). All potential loss of function (LOF) variants (stop gains/losses, splicing and frameshift insertions/deletions) were classified as damaging. All nonsynonymous variants and nonframeshift insertions/deletions were classified as damaging with a CADD score ≥15. Variants were also annotated with and classified as rare based on minor allele frequency (MAF) data from the (i) NHLBI Exome Sequencing Project (European American subjects only; http://evs.gs.washington.edu/EVS/), and (ii) gnomAD (Non-Finnish Europeans only; https://gnomad.broadinstitute.org/).

WES generated genotypes were converted into PLINK format using VCFTOOLS (Danecek et al., 2011), and ROHs were identified using the same method previously described (Simón-Sánchez et al., 2012). Common variation within the exome data of the 28 IPDGC discovery samples were directly compared to the GWAS SNP-chip generated genotypes. The rate of discordant genotypes between platforms for all samples was <5% and all the GWAS-identified ROHs were seen in the exome data. ROHs were then also assessed on UCL exome data from which we identified 60 UCL EOPD samples to include in the discovery phase.

### Candidate variant/gene identification

To maximise efforts to identify novel AR variants within the discovery cohort, it was hypothesised that disease is caused by rare (minor allele frequency [MAF]<0.01) or private (not seen in gnomAD) homozygous potential loss of function (LOF) variants that disrupt protein function and/or stability. The following autosomal variants were therefore prioritised: stop gains/losses, splicing, frameshift insertions/deletions and nonsynonymous variants predicted to be deleterious (CADD≥15) (Kircher et al., 2014). All LOFs were extracted (*i.e*. not just within extended ROH) as it was further hypothesised that additional smaller undetected ROHs may harbour candidate variants. Genes containing prioritised LOFs were taken forward for replication. Due to the absence of phase data, all carriers of two heterozygous variants were assumed to be compound heterozygotes. Where possible, biallelic variants in candidate genes identified in WES analysis were assessed by Sanger sequencing.

### Statistical analyses

Biallelic carriers in each gene were identified and assessed for association with EOPD using logistic regression including the following covariates: coverage metrics, gender and principal components (1-4) for IPDGC WES only. No covariates were available for UCL WES. Fisher’s exact tests were used to assess association with EOPD using combined counts across both replication cohorts. One-sided P-values <0.05 were considered statistically significant. Uncorrected P-values are shown throughout.

### Reagents

The following plasmids were obtained from Addgene: mEmerald-Mito-7 was a gift from Michael Davidson (Addgene plasmid #54160; http://n2t.net/addgene:54160; RRID:Addgene_54160) (Planchon et al., 2011), mito-PAGFP was a gift from Richard Youle (Addgene plasmid #23348; http://n2t.net/addgene:23348; RRID:Addgene_23348) (Karbowski et al., 2004), mApple-TOMM20-N-10 was a gift from Michael Davidson (Addgene plasmid #54955; http://n2t.net/addgene:54955; RRID:Addgene_54955). C-terminal mCherry-tagged MID51 (WT; p.R169W; p.A53V) and C-terminal myc-tagged MID51 (WT; p.R169W; p.A53V) were generated by VectorBuilder. The following reagents were also used: Myc-tag rabbit antibody (Cell Signaling, 2272S) and Myc-Tag (9B11) mouse mAb (Cell Signaling, 2276).

### Cell culture and transfection

HeLa cells (ATCC) were cultured in Dulbecco’s modified Eagle’s medium (DMEM) (Gibco; 11995-065) supplemented with 10% (vol/vol) FBS, 100 units per ml penicillin, and 100μg/ml streptomycin. All cells were maintained at 37°C in a 5% CO^2^ incubator and previously verified by cytochrome *c* oxidase subunit I and short tandem repeat testing, and were tested and found negative for mycoplasma contamination. Cells were transfected using Lipofectamine 2000 (Invitrogen). For live imaging, cells were grown on glass-bottomed culture dishes (MatTek; P35G-1.5-14-C).

### Confocal microscopy

All live cell confocal images were of Hela cells and acquired on a Nikon A1R confocal microscope with GaAsp detectors using a Plan Apo λ 100x 1.45 NA oil immersion objective (Nikon) using NIS-Elements (Nikon). Live cells were imaged in a temperature-controlled chamber (37°C) at 5% CO2 at 1 frame every 2–3s. For mito-PAGFP experiments, the matrix of a subpopulation of mitochondria were selectively labelled by localised photoactivation using a 405nm laser (100% for 4sec) in cells transfected with photoactivatable mitochondrial matrix marker mito-PAGFP and either control (mApple-Tom20) or mCherry MID51 (WT, p.R169W or p.A53V) and fluorescence intensity at a distal region (10µm from the site of photoactivation) was analysed.

### Immunoprecipitation (IP)

To examine MID51 oligomerisation, HeLa cells were transfected for 24h with myc-tagged MID51 (WT, p.R169W or p.A53V) or not transfected (control). Cells were lysed in EBC buffer (Boston Bioproducts; C14-10) with cOmplete*™* Protease Inhibitor Cocktail (Roche; 11873580001) and sonicated. Lysates were immunoprecipitated using Protein G-coupled Dynabeads (Invitrogen; 100003D) incubated in anti-myc antibody (ms), and washed in EBC buffer. Equal total protein levels of immunoprecipitate were analysed by SDS-PAGE and Western blot according to standard protocols using anti-myc antibody (Rb). Measured band intensities of immunoprecipitated Mid51 for oligomers (dimer; tetramer; high molecular weight (HMW)) were normalised to monomeric MID51, and further normalised to MID51 (WT), and expressed as the ratio of MID51 oligomer:monomer for each condition.

### Image analysis

MID51 localisation to mitochondria was quantified as the percentage of cells which had MID51 (mCherry-tagged) colocalised with mitochondria (mEmerald-mito). Mitochondrial fusion dynamics were analysed as the mito-PAGFP fluorescence intensity at a distal region (10µm from the site of photoactivation), and was measured for 5 minutes post-photoactivation, and subsequently normalised to the intensity at t=0 minutes. The mito-PAGFP fluorescence intensity used was the maximum intensity in a 4µmx4µm area in the distal region. Representative traces for mito-PAGFP fluorescence intensity were the average of 3 different cells (from 3 independent experiments) per condition. Immunoblots were quantified using ImageJ (NIH).

### Graphing and figure assembly

Cellular data were analysed using one-way ANOVA with Tukey’s post hoc test (for multiple datasets) or unpaired two-tailed Student *t* test (for two datasets), and statistics are shown comparing MID51 (WT) to MID51 variants. Data presented are means ± s.e.m. All statistical tests were justified as appropriate (see figure legends for details on *n* cells) from *N* ≥ 3 independent experiments (biological replicates) per condition. Statistics and graphing were performed using Prism 7 (GraphPad) software. All videos and images were assembled using ImageJ 1.51j8 (NIH). All final figures were assembled in Illustrator (Adobe).

## Supporting information

ROH_paper_suppl

## Data Availability

The data used is available through application from the following sites:
IPDGC: https://pdgenetics.org/contact
AMP-PD: https://amp-pd.org

## ACKNOWLEDGEMENTS

We would like to thank all of the subjects who donated their time and biological samples to be part of this study. We would also like to thank all members of the International Parkinson’s Disease Genomics Consortium (IPDGC). For a complete overview of members, acknowledgements and funding, please see http://pdgenetics.org/partners.

We would like to thank the Accelerating Medicines Partnership initiative (AMP-PD) for the publicly available whole-genome sequencing data, including cohorts from the BioFIND study, the NINDS Parkinson’s disease Biomarkers Program (PDBP), and MJFF Parkinson’s Progression Marker Initiative (PPMI). As registered users of the AMP-PD, BIB and SJL have access to individual-level data.

Parkinson’s Disease Biomarker Program (PDBP) consortium is supported by the National Institute of Neurological Disorders and Stroke (NINDS) at the National Institutes of Health. A full list of PDBP investigators can be found at https://pdbp.ninds.nih.gov/policy.

Data used in the preparation of this article were obtained from the Parkinson’s Progression Markers Initiative (PPMI) database (www.ppmi-info.org/data). For up-to-date information on the study, visit www.ppmi-info.org. PPMI is sponsored and partially funded by The Michael J. Fox Foundation for Parkinson’s Research (MJFF). Other funding partners include a consortium of industry players, non-profit organizations and private individuals, and include the following (in alphabetical order): Abbvie, Allergen, Amathus Therapeutics, Avid Radiopharmaceuticals, Biogen, BioLegend, Bristol-Myers Squibb, Celgene, Denali, GE Healthcare, Genentech, GlaxoSmithKline, Golub Capital, Handl Therapeutics, Insitro, Janssen Neuroscience, Eli Lilly, Lundbeck, Merck, Meso Scale Discovery, Pfizer, Piramal, Prevail Therapeutics, Roche, Sanofi Genzyme, Servier, Takeda, Teva, UCB, Verily, and Voyager Therapeutics. Industry partners are contributing to PPMI through financial and in-kind donations and are playing a lead role in providing feedback on study parameters through the Partner Scientific Advisory Board (PSAB). Through close interaction with the study, the PSAB is positioned to inform the selection and review of potential progression markers that could be used in clinical testing.

We thank members of the North American Brain Expression Consortium (NABEC) for providing DNA samples derived from brain tissue (dbGaP Study Accession: phs001300.v1.p1). Brain tissue for the NABEC cohort was obtained from the Baltimore Longitudinal Study on Aging at the Johns Hopkins School of Medicine, the NICHD Brain and Tissue Bank for Developmental Disorders at the University of Maryland, the Banner Sun Health Research Institute Brain and Body Donation Program, and from the University of Kentucky Alzheimer’s Disease Center Brain Bank.

